# Hypophosphatemia Is Associated with Post-Operative Ileus After Right Colon Resection

**DOI:** 10.1101/2024.05.11.24307033

**Authors:** Allen T. Yu, Simran Malhotra, Marnie Abeshouse, Esther Yoo, Joseph Sullivan, Alex Huang, Michael C. Plietz, Sergey Khaitov, Alexander J. Greenstein, Patricia Sylla, Sue J. Hahn

**Affiliations:** Icahn School of Medicine at Mount Sinai, New York, NY

**Keywords:** phosphate, post-operative ileus, electrolytes, return of bowel function, colon resection, Crohn’s disease, ileocolic resection

## Abstract

**Background:** Electrolyte imbalances are known to contribute to intestinal ileus. However, the direct impact of hypophosphatemia on post-operative ileus (POI) is unknown.

**Objective:** To describe post-operative phosphate dynamics and if hypophosphatemia is associated with POI after a right colon resection.

**Design:** Comparative retrospective cohort study

**Settings:** High-volume tertiary referral center

**Patients:** Patients who underwent right colon resection, which includes right hemicolectomies and ileocolic resections between 2020 and 2022.

**Main Outcome Measures:** POI incidence, post-surgical phosphate dynamics, and post-operative phosphate deficit and recovery.

**Results:** A total of 396 patients were reviewed, where 68% of resections were for inflammatory bowel disease. Patients had a mean return of bowel function on POD 3.78 ± 1.45. 17.4% of patients overall had POI. Serum phosphate was the most dynamic post-operative electrolyte, with statistically significant differences between POI and non-POI on POD 1, 3, and 7 (*p* < 0.05). Serum phosphate recovery in patients with POI was impaired at 0.11 mg/dL/day versus 0.17 mg/dL/day (*p* < 0.001). Patients with POI had a phosphate deficit that persisted beginning on POD 2, with statistically significant deficits on POD 3-5 (*p* < 0.01), as well as POD 7 (*p* < 0.001). On multivariate analysis, a phosphate deficit on POD 3 (OR_adj_ 9.04, 95% CI 1.38-59.2), POD 5 (OR_adj_ 7.05, 1.13-44.1), and POD 7 (OR_adj_ 47.2, 2.98-749.4) were the only independent risk factors for POI.

**Limitations:** Generalizability of these findings may be limited outside of right colon resections.

**Conclusions:** We have established baseline phosphate dynamics in patients who undergo ileocolic anastomoses. We found POI was associated with a delayed serum phosphate recovery, as well as lower overall phosphate levels. Thus, a potential post-surgical window for intervention with timed phosphate repletion may have the potential to reduce post-operative ileus, need for nasogastric decompression, and ultimately decrease hospital length of stay.

## Introduction

Post-operative ileus (POI), defined as the abnormal slowing or absence of gastrointestinal motility, is a common complication of abdominal surgeries (1, 2). POI typically resolves within 72 hours of the inciting operation and necessitates supportive care (1, 3). However, prolonged POI, lasting greater than 72 hours, contributes to a protracted hospital stay and results in delayed enteral feeding (4). The poor nutritional intake can lead to subsequent impaired wound healing and increased infection susceptibility (2, 4). Longer hospitalizations due to POI financially burdens the health system, estimated to cost 750 million dollars annually in the United States (2, 5).

While the etiology of prolonged POI is multifactorial, a well-studied mechanism is the effect of electrolyte derangements on gut motility (2, 6). Multiple studies have shown that hypokalemia, hypocalcemia, and hyponatremia are each associated with POI (6, 7), but few studies have focused on the effect of hypophosphatemia on POI. The phosphate ion is ubiquitously required in many cellular processes, including signaling, modification of proteins and nucleic acids, and cellular energetics. At the molecular point of view, smooth muscle that propagates peristalsis is due to the contraction of myosin heads while bound to actin microfilaments through the hydrolysis of adenosine triphosphate (ATP) to adenosine diphosphate (ADP). Thus, it is possible that hypophosphatemia would cause a deficit of ATP production and cause POI.

Hypophosphatemia, defined as a serum phosphate level < 2.5 mg/dL, has been highlighted in patients who have undergone gastrointestinal resections, including hepatic, pancreatic, colorectal, and gastric. These studies have shown that low levels of phosphate following post-operative day (POD) 2 are associated with increased rates of complications and morbidity (8–10). One study concluded that hypophosphatemia may have some prognostic value for abdominal organ specific surgeries, possibly serving as an early indication for anastomotic leak (9). Another study showed hypophosphatemia during POD 5 through 7 after cytoreductive surgery or hyperthermic intraperitoneal chemotherapy is associated with more complications and risk of anastomotic leak (8).

A gap in the literature exists when exploring the effect of hypophosphatemia on POI as well as a description of post-surgical electrolyte dynamics. However, no standard procedure exists to study this phenomenon and different surgeries can cause different rates of return of bowel function. A right colon resection with an ileocolic anastomosis is a typically well-tolerated anastomosis with a length of hospital stay that is within the effect range of our study (11–13). It is a standard surgical procedure used to treat small bowel pathologies, most commonly Crohn’s disease, and ascending colon cancer (11, 14). Here, we aimed to explore the dynamics of post-surgical electrolyte shifts with a focus on serum phosphate, as well as investigate hypophosphatemia’s influence on POI in a patient population who received a right colon resection.

## Methods

A retrospective study was performed on patients who underwent a right colon resection at a tertiary referral center, largely identified using current procedural terminology codes: 44160 and 44205, between the dates of January 1^st^, 2020 and January 1^st^, 2022. This included patients who underwent right hemicolectomies or ileocolic resections where an ileocolic anastomosis was made. Patients > 18 years old with phosphate labs and hospital notes where bowel function was recorded were included. Patients with end stage renal disease, need for dialysis, ostomy creation, and with a past medical history of known hyperphosphatemia were excluded.

Perioperative serum electrolyte levels including sodium potassium, calcium, magnesium, and phosphate were recorded up to POD 7. Return of bowel function was defined as the day the patient passed flatus. POI was defined as having return of bowel function on a POD one standard deviation above the mean (POD5). Patient demographics, length of stay after surgery, and comorbidities were compared between patients with or without POI. Students T-tests and Mann-Whitney U-tests were used for comparative statistics, and linear regressions were performed to determine rate of change. Phosphate deficit was calculated from POD 1 lab values as a baseline. Multivariate binary logistic regressions were performed with the outcome variable as POI or nasogastric tube usage, with covariates being salient clinical demographics as well as variables significant on univariate analysis. STROBE reporting guidelines were followed.

## Results

### Patient population and definition of post-operative ileus

A total of 396 patients were found to have met the inclusion criteria, with a median age of 44 years, 53.5% male, and median BMI of 23.6. Most patients were ASA II (230 patients, 58.1%), and ASA III (142, 35.9%). 21% of patients had hypertension, 7.8% had diabetes, 6.1% had coronary artery disease, and 2.0% had chronic obstructive pulmonary disorder. The most common indication for a right colon resection is inflammatory bowel disease (271, 68.4%), and the second most common indication was malignancy (107, 27.0%). The majority of cases were performed via the laparoscopic approach (309, 78.0%), and 87 (22.0%) were performed open. 64 (16.2%) patients had an abscess that was concurrently drained, and 23 (5.8%) required insertion of a nasogastric tube due to oral intolerance. The vast majority of cases were elective at 96.2%, and only 15 (3.8%) were emergent. The mean POD that flatus, or return of bowel function by our definition, was 3.46 ± 1.25. While not all patients had a bowel movement during their admission, of those who did (292 patients, 73.7%), the mean POD was 3.78 ± 1.45. The mean length of stay after surgery was 5.11 ± 2.78 days. Post-operative pain regimen differed among patients, but we categorized them into three general categories: non-opioid multimodal pain control, opioid-based pain control as needed, and patient controlled analgesia. 349 (88.1%) of patients had patient controlled analgesia in our population, followed by 28 (7.1%) who had a non-opioid multimodal pain regimen (Table 1).

**Table 1.** Patient demographics.

| Characteristic | N = 396 | %/IQR/SD |
| --- | --- | --- |
| Age (years, median, IQR) | 44.0 | 29.0-64.3 |
| Sex (%) |  |  |
| Male | 212 | 53.5 |
| Female | 182 | 46.0 |
| Non-Binary | 2 | 0.5 |
| Body Mass Index (kg/m<sup>2</sup>, median, IQR) | 23.6 | 20.7-28.1 |
| ASA (%) |  |  |
| I | 4 | 1.0 |
| II | 230 | 58.1 |
| III | 142 | 35.9 |
| IV | 20 | 5.1 |
| Past Medical History (%) |  |  |
| Hypertension | 83 | 21.0 |
| Coronary artery disease | 24 | 6.1 |
| Diabetes mellitus | 31 | 7.8 |
| Chronic obstructive pulmonary disorder | 8 | 2.0 |
| Indication (%) |  |  |
| Malignancy | 107 | 27.0 |
| Inflammatory bowel disease | 271 | 68.4 |
| Other | 18 | 4.5 |
| Status (%) |  |  |
| Elective | 381 | 96.2 |
| Emergent | 15 | 3.8 |
| Approach (%) |  |  |
| Laparoscopic | 309 | 78.0 |
| Open | 87 | 22.0 |
| Abscess (%) | 64 | 16.2 |
| Nasogastric tube (%) | 23 | 5.8 |
| POD for first flatus (mean, SD) | 3.46 | 1.25 |
| POD for first bowel movement (mean, SD) | 3.78 | 1.45 |
| Length of Stay (mean, SD) | 5.11 | 2.78 |
| Pain regimen (%) |  |  |
| Non-opioid multimodal | 28 | 7.1 |
| Opioid PRN | 19 | 4.8 |
| Patient controlled analgesia | 349 | 88.1 |

From here, we graphed the distribution of return of bowel function in our patients (Figure 1). To determine a cut-off for our definition of POI, we empirically established that one standard deviation above the mean day for return of bowel function to be POI. Thus, in our population, 69 patients (17.4%) had our definition of post-operative ileus.

**Figure 1.**
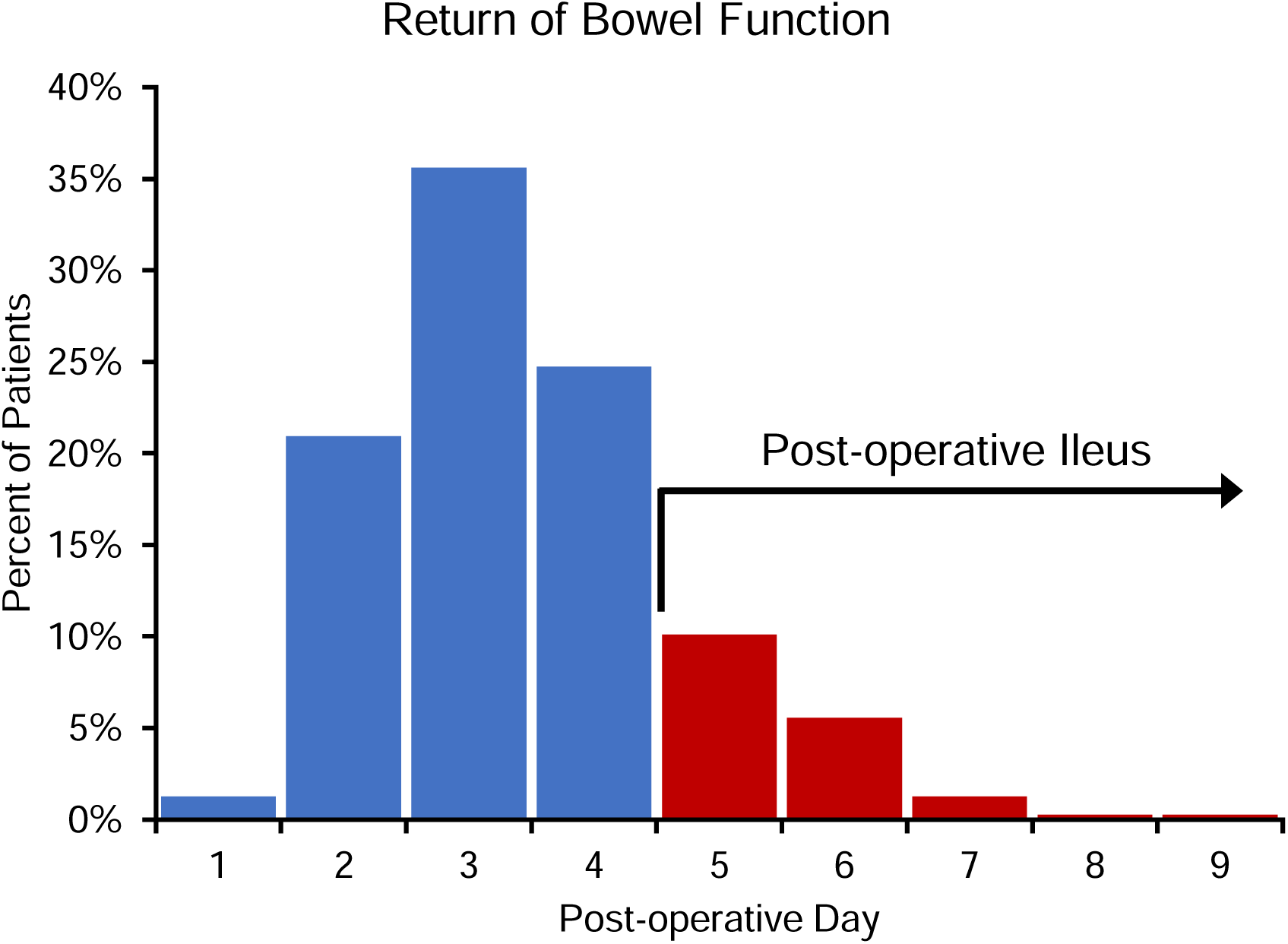
Distribution of flatus and post-operative day

### Stratification of patients by post-operative ileus

The included patients were stratified to those with POI and those without POI. Patients with POI were significantly older (55.0 years vs 42.0, *p* < 0.01), had a higher ASA classification (*p* < 0.01), and had a higher incidence of hypertension (30.4% vs 19.0%, *p* < 0.05), diabetes mellitus (14.5% vs 6.4%, *p* < 0.05), and chronic obstructive pulmonary disorder (5.8% vs 1.2%, *p* < 0.05). Patients with POI were more likely to have an open approach (39.1% vs 18.3%, *p* < 0.001). Sex, body mass index (BMI), history of coronary artery disease, and presence of an abscess, were not significant between the two groups (Table 2).

**Table 2.** Stratification of population by prolonged post-operative ileus.

| **Characteristic**<br>**N = 396** | No Ileus<br>N = 327 | Ileus<br>N = 69 | p-value |
| --- | --- | --- | --- |
| Age (median, IQR) | 42.0 (28.0-62.0) | 55.0 (35.0-74.0) | **0.002** |
| Sex (%) |  |  | 0.151 |
| Male | 168 (51.4) | 44 (63.8) |  |
| Female | 157 (48.0) | 25 (36.2) |  |
| Non-Binary | 2 (0.6) | 0 (0.0) |  |
| Body Mass Index (kg/m<sup>2</sup>) | 23.8 (20.7-28.5) | 23.0 (20.4-25.9) | 0.224 |
| ASA (%) |  |  | **0.006** |
| I | 3 (0.9) | 1 (1.4) |  |
| II | 197 (60.2) | 33 (47.8) |  |
| III | 116 (35.5) | 26 (37.7) |  |
| IV | 11 (3.4) | 9 (13.0) |  |
| Past Medical History (%) |  |  |  |
| Hypertension | 62 (19.0) | 21 (30.4) | **0.049** |
| Coronary artery disease | 18 (5.5) | 6 (8.7) | 0.464 |
| Diabetes mellitus | 21 (6.4) | 10 (14.5) | **0.043** |
| Chronic obstructive pulmonary disorder | 4 (1.2) | 4 (5.8) | **0.047** |
| Indication (%) |  |  | **0.028** |
| Malignancy | 86 (26.3) | 21 (30.4) |  |
| Inflammatory bowel disease | 230 (70.3) | 41 (59.4) |  |
| Other | 11 (3.4) | 7 (10.1) |  |
| Status (%) |  |  | 1.000 |
| Elective | 315 (96.3) | 66 (95.7) |  |
| Emergent | 12 (3.7) | 3 (4.3) |  |
| Approach (%) |  |  | **<0.001** |
| Laparoscopic | 267 (81.7) | 42 (60.9) |  |
| Open | 60 (18.3) | 27 (39.1) |  |
| Abscess | 53 (16.2) | 11 (15.9) | 1.000 |
| Nasogastric tube | 11 (3.4) | 12 (17.4) | **<0.001** |
| Flatus on POD | 3.02 ± 0.79 | 5.57 ± 0.81 | **<0.001** |
| Bowel movement on POD | 3.36 ± 1.18 | 5.64 ± 1.06 | **<0.001** |
| Length of Stay | 4.47 ± 1.69 | 8.16 ± 4.43 | **<0.001** |
| Pain regimen |  |  | 0.358 |
| Non-opioid multimodal | 23 (7.0) | 5 (7.2) |  |
| Opioid PRN | 18 (5.5) | 1 (1.4) |  |
| Patient controlled analgesia | 286 (87.5) | 63 (91.3) |  |

Post-operatively, patients with POI were more likely to have a nasogastric tube inserted (17.4% vs 3.4%, *p* < 0.001). As we stratified these patients by POI, it is expected to see that patients with POI had a longer time to passing flatus (POD 5.57 vs 3.02, *p* < 0.001), having a bowel movement (POD 5.64 vs 3.36, *p* < 0.001), and length of stay after surgery (8.16 days vs 4.47, *p* < 0.001). There was no statistically significant difference in post-operative pain regimen between the two groups (Table 2).

### Analysis of electrolyte dynamics

Serum levels of the most common electrolytes were collected and their trended. The general trend of serum phosphate is striking, given its drastic drop that’s greater than 1 mg/dL (25%) on POD 2, and then recovery over the next post-operative hospital course (Figure 2A). Serum sodium and calcium both have a relatively stable post-operative course (Figure 2B and 2E), while serum potassium and magnesium have a gentle decrease over the seven recorded post-operative days (Figure 2C and 2D).

**Figure 2.**
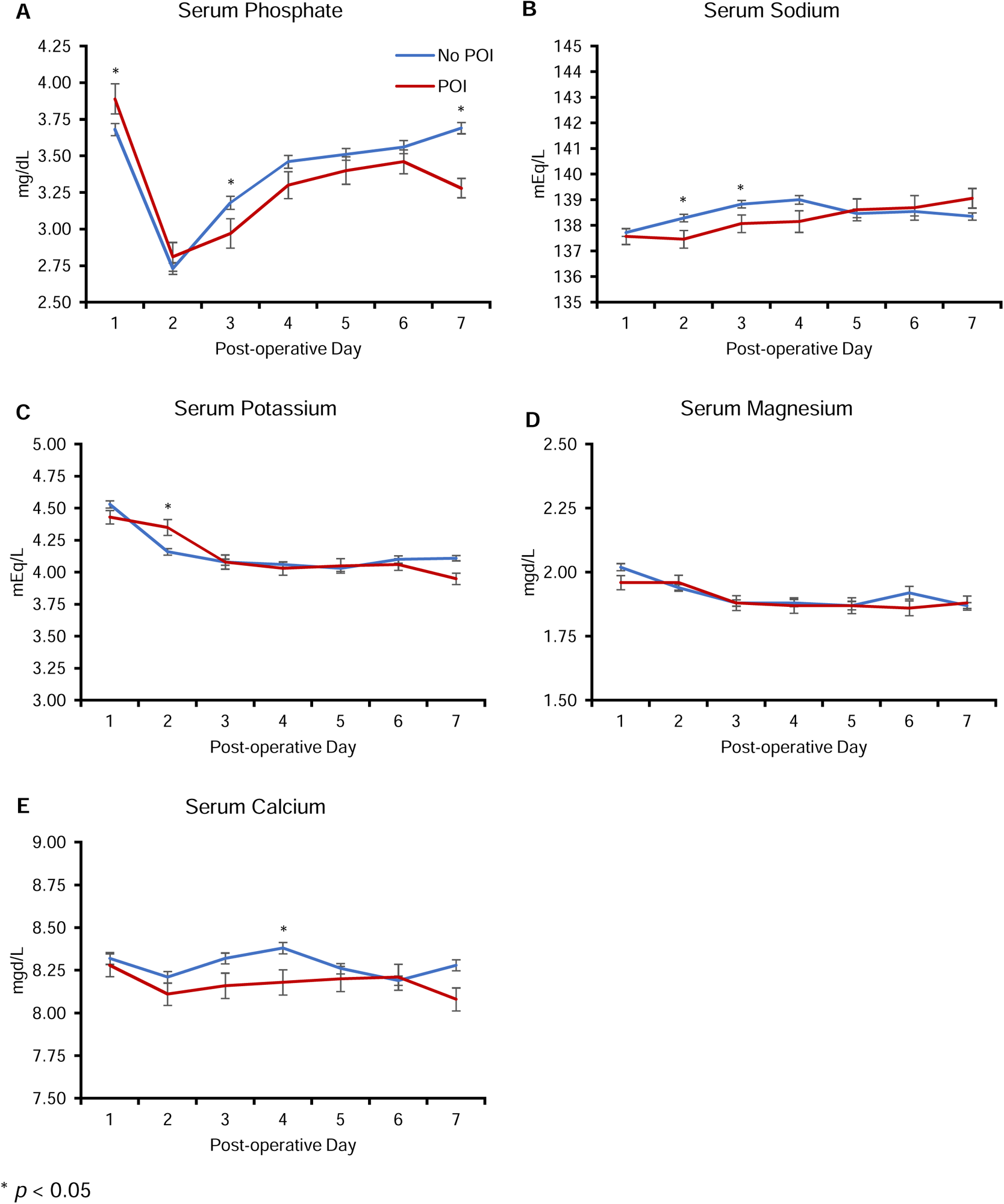
Electrolyte dynamics over time stratified by post-operative ileus

These electrolyte profiles were then separated into our two populations, those with POI and those without. Patients with POI were observed to have a statistically significant higher POD 1 serum phosphate, and then slowed recovery on POD 3, and finally plateauing of recovery on POD 7, but never reaching baseline (Figure 2A, *p* < 0.05). Serum sodium had significant differences on POD 2 and POD 3, but normalized between the two groups thereafter (Figure 2B). Patients with POI had a higher POD 2 serum potassium by 0.19 mEq/L (*p* < 0.05), but also normalized on POD 3 (Figure 2C). Serum calcium was significantly lower on POD 4 in patients with POI, but was otherwise not significant during the patient’s hospital stay (Figure 2E). Serum magnesium as not statistically significant between the two groups (Figure 2D).

### Serum phosphate recovery and deficit

Given the highly dynamic state of serum phosphate in the post-surgical period, an in-depth analysis in the recovery and deficit of serum phosphate was performed. While serum phosphate drops on POD 2, this is apparent in all patients. However, there was a slower rate of serum phosphate recovery in patients with POI than those without. Linear regressions showed that the recovery rate of phosphate in POI patients was 0.11 mg/dL per day vs 0.17 mg/dL per day (*p* < 0.001). This is roughly a 33% decrease in the rate of phosphate recovery (Figure 3A).

**Figure 3.**
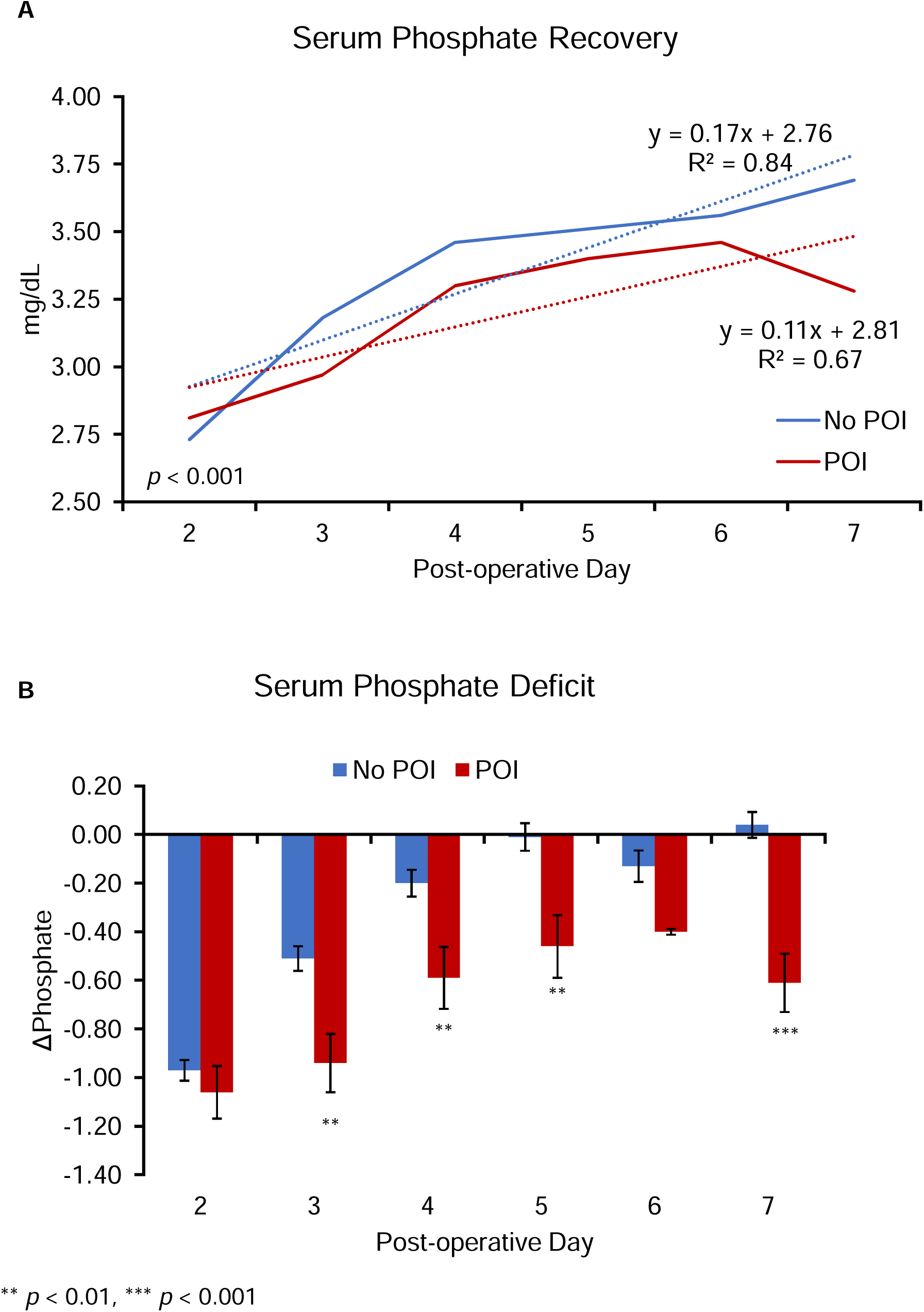
Phosphate deficit and recovery

As patients had different baseline serum phosphates, we calculated individual patients’ serum phosphate deficit on each post-operative day, with POD 1 serum phosphate as the baseline. We found no difference in deficit on POD 2, but statistically significant deficits on POD 3-5 (*p* < 0.01), as well as POD 7 (*p* < 0.001, Figure 3B). Patients without POI largely have no phosphate deficit by POD 5. This is consistent with our previous analyses where there is a lower serum phosphate on POD 3, and a slower recovery over the hospital course.

To determine if phosphate deficits were independent risk factors for POI, a multivariate analysis was performed with POI as the outcome variable. Clinically relevant covariates such as age, sex, and pain regimen were included, as well as those which were significant on univariate analysis (ASA, hypertension, diabetes, indication, approach). Phosphate deficits (ΔPhosphate) found to be significant based on Figure 2A (POD 3, 4, 5, and 7) were included as well. Interestingly, the only statistically significant independent risk factors for POI was a phosphate deficit on POD 3 (OR_adj_ 9.04, 95% CI 1.38-59.2, *p* = 0.019), POD 5 (OR_adj_ 7.05, 1.13-44.1, *p* = 0.031), and POD 7 (OR_adj_ 47.2, 2.98-749.4, *p* = 0.006) (Table 3).

**Table 3.** Multivariate analysis of post-operative ileus.

| **Post-operative Ileus**<br>**N = 69** | **OR<sub>adj</sub> (95% CI)** | **p-value** |
| --- | --- | --- |
| Age | 1.02 (0.97-1.08) | 0.448 |
| Sex (%) |  |  |
| Male | 1.0 (ref) |  |
| Female | 1.53 (0.21-11.26) | 0.371 |
| ASA (%) |  |  |
| II | 1.0 (ref) |  |
| III | 5.05 (0.57-44.6) | 0.167 |
| IV | 4.68 (0.17-126.6) | 0.325 |
| Past Medical History |  |  |
| Hypertension | 0.59 (0.09-3.99) | 0.314 |
| Diabetes mellitus | 9.75 (0.50-190.0) | 0.107 |
| Indication (%) |  |  |
| Malignancy | 1.0 (ref) |  |
| Inflammatory bowel disease | 0.87 (0.06-12.9) | 0.877 |
| Other | 1.60 (0.02-119.9) | 0.815 |
| Approach (%) |  |  |
| Laparoscopic | 1.0 (ref) |  |
| Open | 3.03 (0.41-22.5) | 0.166 |
| Pain regimen |  |  |
| Non-opioid multimodal | 1.0 (ref) |  |
| Patient controlled analgesia | 6.60 (0.09-472.9) | 0.291 |
| ΔPhosphate | | |
| ΔPhos3 | 9.04 (1.38-59.2) | **0.019** |
| ΔPhos4 | 0.49 (0.11-2.25) | 0.694 |
| ΔPhos5 | 7.05 (1.13-44.1) | **0.031** |
| ΔPhos7 | 47.2 (2.98-749.4) | **0.006** |

### Sub-analysis of patients required nasogastric tubes

As insertion of a nasogastric tube generally indicates clinical POI, sub-analysis was performed of the patients who had a nasogastric tube to determine which risk factors contributed to the necessity of placing one. On univariate analysis, patients who were older (58.0 years, *p* = 0.002), male (78.3%, *p* = 0.049), higher ASA classification (*p* = 0.009), indication of malignancy (34.8%, p = 0.004), and taken an open approach (82.6%, *p* < 0.001), all had a statistically significant difference between patients who had a nasogastric tube placed and those who did not. Post-operatively, these patients also had delayed flatus on POD 4.78 (*p* < 0.001), delayed bowel movements on POD 5.57 (*p* < 0.001), and increased length of stay to 10.0 days (*p* < 0.001). Also as expected, these patients were significantly enriched for our definition of POI (52.2%, *p* < 0.001). Only a phosphate deficit on POD 4 was statistically significant (-0.81 mg/dL, *p* = 0.020).

Multivariate analysis with the significant variables as covariates demonstrated that only the open approach and having a delayed flatus were significant risk factors for requiring a post-operative nasogastric tube (OR_adj_ 18.2, 4.42-75.0, *p* < 0.001, and OR_adj_ 2.46, 1.15-5.29, *p* = 0.021, respectively). Phosphate deficient on POD 4 was trending towards significance (OR_adj_ 1.64, 0.9-2.96, *p* = 0.10). All other variables were not independent risk factors for post-operative nasogastric tube placement (Table 4).

**Table 4.** Sub-analysis of patients requiring a post-operative nasogastric tube.

| **Nasogastric tube**<br>**n = 23** | **n (%) / mean ± SD** | **p-value** | **Multivariate**<br>**OR (95% CI)** | **p-value** |
| --- | --- | --- | --- | --- |
| Age (median, IQR) | 58.0 (46.5-74.5) | **0.002** | 0.98 (0.94-1.03) | 0.449 |
| Sex |  | **0.049** |  |  |
| Male | 18 (78.3) |  | 1.0 (Ref) |  |
| Female | 5 (21.7) |  | 0.23 (0.01-1.11) | 0.067 |
| Body Mass Index (kg/m<sup>2</sup>) | 24.8 (22.1-28.9) | 0.150 |  |  |
| ASA |  | **0.009** |  |  |
| I | 1 (4.3) |  | 1.0 (Ref) |  |
| II | 9 (39.1) |  | 0.36 (0.01-15.1) | 0.589 |
| III | 9 (39.1) |  | 0.44 (0.01-18.8) | 0.667 |
| IV | 4 (17.4) |  | 0.64 (0.01-43.9) | 0.836 |
| Past Medical History (%) |  |  |  |  |
| Hypertension | 8 (34.8) | 0.162 |  |  |
| Coronary artery disease | 1 (4.3) | 1.000 |  |  |
| Diabetes mellitus | 1 (4.3) | 0.805 |  |  |
| Chronic obstructive pulmonary disorder | 0 (0.0) | 1.000 |  |  |
| Indication (%) |  | **0.004** |  |  |
| Malignancy | 8 (34.8) |  | 1.0 (Ref) |  |
| Inflammatory bowel disease | 11 (47.8) |  | 0.22 (0.03-1.35) | 0.103 |
| Other | 4 (17.4) |  | 1.11 (0.11-11.4) | 0.931 |
| Approach (%) |  | **<0.001** |  |  |
| Laparoscopic | 4 (17.4) |  | 1.0 (Ref) |  |
| Open | 19 (82.6) |  | 18.2 (4.42-75.0) | **<0.001** |
| Abscess | 5 (21.7) | 0.630 |  |  |
| Flatus on POD | 4.78 ± 1.57 | **<0.001** | 2.46 (1.15-5.29) | **0.021** |
| Bowel movement on POD | 5.57 ± 1.90 | **<0.001** | | |
| Length of Stay | 10.0 ± 6.83 | **<0.001** | | |
| Pain regimen |  | 0.450 |  |  |
| Non-opioid multimodal | 1 (4.3) |  |  |  |
| Opioid PRN | 0 (0.0) |  |  |  |
| Patient controlled analgesia | 22 (95.7) |  |  |  |
| Post-operative Ileus | 12 (52.2) | **<0.001** | 0.41 (0.06-2.97) | 0.376 |
| ΔPhosphate | | | | |
| ΔPhos3 | -0.90 ± 1.14 | 0.124 | | |
| ΔPhos4 | -0.81 ± 1.26 | **0.020** | 1.64 (0.9-2.96) | 0.104 |
| ΔPhos5 | -0.57 ± 0.96 | 0.090 | | |
| ΔPhos6 | -0.44 ± 1.16 | 0.491 | | |
| ΔPhos7 | -0.57 ± 1.27 | 0.302 | | |

## Discussion

In this study, we explored the post-operative dynamics of serum electrolytes, particularly serum phosphate, in a population who received a right colon resection and its relation to post-operative ileus. We empirically defined POI in our population as one standard deviation above the mean POD where the population had flatus. We found that serum phosphate has a drastic decrease on POD 2, and those with POI had an impaired recovery of serum phosphate and increased phosphate deficit. On multivariate analysis, a large phosphate deficit on POD 3 was an independent risk factor for having POI, with an OR of 9.04, and a minimum effect size of 1.38. These results imply that there is a window for intervention, where post-operative patients can receive standard serum phosphate repletions in preparation for the drop seen in every patient on POD 2, to encourage ATP production and drive peristalsis, to prevent the phosphate deficit seen on POD 3. There is precedent to drive return of bowel function with phosphate repletions up to 5.5 mg/dL in a patient with myogenic ileus (15).

Definitions of post-operative ileus differ in the literature, and we settled upon our definition empirically, based on our population data. Our rate of POI of 17.4% is very comparable to the existing literature of 10.2% to 17.2% (7, 16, 17). We found this definition to be more stringent as some patients would receive a nasogastric tube in the early post-operative period, and may still have return on bowel function < POD 5. In addition, in our population, patients are placed on a clear liquid diet and can advance as tolerated ad lib. Thus, many are not placed NPO and are self-regulated. Another POI scoring system, the I-FEED classification, subdivides patients into normal, postoperative GI intolerance (POGI), and postoperative GI dysfunction (POGD) (18). Many of our patients with delayed return of bowel function typically did not exhibit significant vomiting or medication resistant nausea, and thus would not be classified as POGI or POGD. We believe our criteria of having flatus > POD 5 is able to capture more patients for analysis in the POI category.

On univariate analysis, patients who were older, higher ASA classification, with a past medical history of hypertension, diabetes, or chronic obstructive pulmonary disorder, and the open approach, were more likely to have POI. These factors are in-line with previous studies who also found similar risk factors for POI (16, 19). In our study, when the covariate of phosphate deficit was included in the multivariate analysis, the previous factors were no longer significant due to the large effect size that serum phosphate has on POI. Thus, we believe that pre-emptively repleting serum phosphate will help decrease the incidence of POI.

At our institution and others, it is standard practice to replete serum phosphate with oral neutral phosphate, or intravenous sodium or potassium phosphate if it is lower than 2.5 or 3.0 mg/dL (20). Our patients all underwent standard phosphate repletions; however, the vast majority of patients were not below 2.5 mg/dL (Figure 2A). While we replete phosphate with 15 mEq if it is below 3.0 mg/dL, we still see a decreased recovery in our POI patients on POD 3. The repletions on POD 2 are reactive, and thus the bowel possibly has already entered a state of depleted ATP (21). Once the patient’s serum phosphate is above 3.0 mg/dL, repletions are no longer given per standard operating protocols. As emphasized in numerous figures in this paper, we believe repletions should be given earlier and to a higher level, as it is important to replete the patient back to their original baseline, rather than an absolute value. Interestingly, the incidence of POI in patients with a baseline serum phosphate level > 5.0 mg/dL is 40.9%, more than two-fold higher than our population average of 17.4%.

A potential limitation is our choice of a right colon resection as a standard procedure to study POI and phosphate dynamics. Ideally, the chosen procedure would be of minimal risk, similar to a diagnostic laparoscopy. However, the LOS is too short to obtain serum phosphate levels or wait for return of bowel function. Bariatric patients typically have a length of day of 1 to 2 days, and are also not within the resolution to capture electrolyte dynamics (22). POI in colectomies have also been well studied, but have an average LOS of 2 to 3 days as well, with some institutions performing same day colectomies (23). Thus, a right colon resection is theoretically able to capture a wide range of ages, indications, and timing of return of bowel function. These patients are an otherwise relatively healthy population, and would not have undergone chemotherapy. The potential drawback of this at our institution is the high volume of inflammatory bowel disease patients, which may not be representative of others. Interestingly, the presence of an abscess did not affect POI, as the bowel being anastomosed in patients with inflammatory bowel disease is inherently more abnormal than the bowel in malignancy.

To our knowledge, we are the first to associate hypophosphatemia with post-operative ileus at this temporal resolution, and describe the post-operative dynamics of serum phosphate. We find that having a phosphate deficit an independent risk factor for having POI greater than POD 5. This opens up a window for an inexpensive and accessible post-operative intervention. Future studies should explore aptly timed infusions of phosphate to reduce the incidence of post-operative ileus, and ultimately decrease hospital length of stay.

## Data Availability

All data produced in the present study are available upon reasonable request to the authors

## Acknowledgements

The authors thank the members of the Department of Surgery at the Mount Sinai Hospital System for their input in the project. We also thank the American Society for Colon and Rectal Surgeons for accepting this a post presentation, presented on June 4^th^, 2023 at Seattle, WA.

## Availability of data and materials

The datasets used and/or analyzed during the current study are available from the corresponding author on reasonable request.

## Ethics approval and consent to participate

The study was IRB-approved: IRB# STUDY-22-00711

